# The causal effect of cigarette smoking on healthcare costs

**DOI:** 10.1101/2022.07.05.22277228

**Authors:** Padraig Dixon, Hannah Sallis, Marcus Munafo, George Davey Smith, Laura Howe

## Abstract

Knowledge of the impact of smoking on healthcare costs is important for establishing the external effects of smoking and for evaluating policies intended to modify this behavior. Conventional analysis of this association is difficult because of omitted variable bias, reverse causality, and measurement error. We approached these challenges using a Mendelian Randomization study design, in which genetic variants associated with smoking behaviors were used as instrumental variables. We undertook genome wide association studies to identify genetic variants associated with smoking initiation and a composite index of lifetime smoking on up to 300,045 individuals in the UK Biobank cohort. These variants were used in two-stage least square models and a variety of sensitivity analyses. All results were concordant in indicating a substantial impact of each smoking exposure on annual inpatient hospital costs Our results indicate a substantial impact of smoking on hospital costs. Genetic liability to initiate smoking – ever versus never having smoked – was estimated to increase mean per-patient annual hospital costs by £477 (95% confidence interval (CI): £187 to £766). A one unit change in genetic liability a composite index reflecting the cumulative health impacts of smoking was estimated to increase these costs by £204 (95% CI: £105 to £303). Models conditioning on the causal effect of risk tolerance were not robust to weak instruments for this exposure. Our findings have implications for the scale of external effects that smokers impose on others, and on the probable cost-effectiveness of smoking interventions.

## 1 Introduction

Over one billion people smoked tobacco products – mostly cigarettes – in 2015 [1]. Despite declines in prevalence in many industrialized countries [1-3], smoking continues to be associated with substantial morbidity and mortality [4, 5]. Smoking is arguably the most damaging of all voluntary health behaviors [6, 7] and is associated with a variety of adverse economic and socioeconomic outcomes [8]. Here we study the causal association of smoking with one such economic outcome: healthcare costs.

The association between smoking and healthcare costs is important [7]. Accurate estimates of the healthcare expenditure attributable to smoking are necessary to calculate the external effects associated with smoking, and these externalities inform and underpin many government interventions intended to prevent smoking [9-12]. Causal evidence on the effect of smoking on healthcare costs is also necessary for the robust evaluation of specific interventions that aim to prevent smoking and to treat its downstream consequences. Decision-making by individual smokers may be improved with better information about the non-health consequences of smoking [13].

However, establishing the causal effect of cigarette smoking on healthcare costs is challenging [7, 14]. Observed associations of smoking with healthcare costs may arise because smoking is indeed a cause of healthcare costs, because smoking is itself partly determined by healthcare costs, or because smoking is associated with causes or consequences of processes that influence healthcare costs. In general, it is not clear if the factors that may predispose an individual to smoke are themselves independent determinants of healthcare costs. For example, smoking tends to cluster with other behaviors known or suspected to affect healthcare costs, including high body mass index (BMI), poor diet, alcohol consumption and low physical activity [15-19]. Smoking is also heavily socially patterned, and lower socio-economic status groups are more likely to initiate and continue smoking [20, 21]. These groups may have access to fewer resources to ameliorate the consequences of incident disease and its progression [22] [23, 24].

Smoking is also more prevalent amongst groups defined by health status. For example, smoking is more common amongst individuals with depression and schizophrenia [25, 26] [27, 28].

These associations may affect all or some of smoking initiation, smoking intensity, and smoking cessation. For example, smoking influences disease incidence (such as lung cancer) which in turn may prompt cessation. Smoking may reflect elements of self-medication [29] or a desire to control one’s weight [30]. Smoking may therefore be both a cause and consequence of health status and other circumstances [14, 31].

Analysis of the effects of smoking may also be complicated by measurement error, including from inaccurate recollection of smoking history. More subtly, even if declarations of smoking habits are accurate, self-reported smoking patterns (“two packs a day for fifteen years”) may not fully capture the consequences of smoking on health. Instead, the cumulative physiological insult of lifetime smoking (reflecting both duration and intensity) on respiratory and other functions may be a better measure for this exposure.

Measured associations of smoking with healthcare cost may also partly reflect wider attitudes to risk tolerance, including impulsivity and behavioral disinhibition. In the Grossman model of health capital accumulation [32], differences between individuals in attitudes to risky behaviors such as smoking may reflect differences in risk attitudes. While empirical support for between-individual differences of this type in line with the Grossman model is sparse [33], it does point to the empirical challenges of distinguishing between smoking and risk tolerance in causal analyses.

These difficulties with robust causal inference comprise the classic issues of omitted variable bias, simultaneity and measurement error [7]. We used Mendelian Randomization to address these issues. Using Mendelian Randomization, and subject to the assumptions of instrumental variable analysis, causal effects of smoking on healthcare cost can be robustly identified by perturbations to genetic variants associated with liability to smoke cigarettes. Since this genetic variation is fixed at conception, it cannot be affected by reverse causation. By virtue of the quasi-random allocation of genetic variation at conception, it will be independent of many confounding variables that might otherwise affect the association between smoking and healthcare cost. We attempted to account for measurement error by using a newly developed index of lifetime smoking that reflects the impact of both smoking duration (encompassing initiation and cessation) and smoking intensity on mortality and health.

## 2 Genetic variants as instrumental variables

We very briefly review the requirements for valid instrumental variable analysis in the context of Mendelian Randomization. Many introductions are available [34-37] and a more extended discussion accompanied by directed acyclic graphs is provided in supplementary material.

An allele refers to the specific genetic code at particular location or locus in the genome. Individuals have two alleles at each location, one inherited from each parent according to Mendel’s first law (the random segregation of alleles) and second law (independent assortment of alleles for different traits) of inheritance. Single nucleotide polymorphisms (SNPs) are examples of genetic variation subject to Mendel’s first and second laws. SNPs are single changes to the nucleotides that make up the genomic code. Some SNPs are known to be associated with disease, traits, or specific behaviors. These associations are usually obtained using genome-wide association studies (GWASs). A variety of different SNPs have been found to robustly associate with various smoking phenotypes.

For example, the *CHRNA5–A3–B4* genomic locus contains the rs16969968 SNP, which is associated with the heaviness of smoking. Individuals with two copies of the less common allele (the “AA” genotype) of this SNP increase the amount of smoke inhaled when nicotine content in cigarettes is increased, whereas individuals with two copies of the more common or major allele do not exhibit this compensatory behavior [14]. Each copy of this allele amounts to approximately one additional cigarette per day in smokers [38]. The *CHRNA5–A3–*B4 locus was identified in various GWAS of different diseases relating to smoking including chronic obstructive pulmonary disease and it is associated both with an earlier diagnosis and an increased risk of lung cancer [39]. It is now apparent that smoking is responsible for the association of this locus with smoking-related diseases.

These associations of SNPs with disease, and their conditionally random allocation at concept, establish the potential for this form of genetic variation to be used as instrumental variables in causal analysis. Briefly, variants should affect the exposure of interest, and have no effects on the outcome (healthcare costs in this case) that are not mediated via its influence on the exposure. The latter assumption embodies two further assumptions – first that the variant does not influence confounders the exposure and the outcome (sometimes referred to as correlated pleiotropy), and second that the variant influences the outcome only via the exposure and not through channels that are independent of the exposure (sometimes referred to as uncorrelated pleiotropy).

Correlated pleiotropy may arise when a SNP affects more than one trait through a shared heritable factor and may lead to false positives when assessing the association between the genetic instruments we use for smoking and risk phenotypes; that is, the appearance of a causal relationship when none may exist in truth. For example, a SNP may influence healthcare costs by its influence on smoking but also via its influence on risk attitudes.

Uncorrelated pleiotropy will arise if SNPs are correlated with other SNPs that themselves influence the outcome independently of the exposure of interest. This correlation arises as a consequence of linkage disequilibrium, which typically occurs when variants located in close physical proximity on the genome are inherited together [40]. SNPs may also influence more than one outcome phenotype through independent pathways, and this will also give rise to this type of pleiotropy.

We expand on these potential violations of the instrumental variable assumptions in supplementary material. We describe below in the Methods section how we assessed and accounted for their presence.

## 3 Methods

### 3.1 Conventional multivariable analysis

We implemented conventional multivariable linear regression as well as Mendelian Randomization. The conventional linear regression models related healthcare cost to both smoking status exposures, controlling in each case for age, sex, and Biobank recruitment centre. We implemented this minimally-adjusted model as a basis for comparison with the instrumental variable models.

### 3.2 Instrumental variable analysis

Instrumental variables were calculated using versions of the Wald ratio. In Mendelian Randomization, the Wald ratio is calculated as the ratio of the association between the SNP(s) and outcome to the association between the SNPs and the exposure [41]. Denoting the SNP-exposure association as *β*_*exp*_, and the SNP-outcome association as *β*_*out*_, the Wald ratio for the causal instrumental variable estimate is

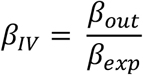

This ratio is equivalent to the two-stage least squares (2SLS) model when a single instrument is used. The first stage of a 2SLS regresses an exposure variable on the instrument variables. The predicted values of the smoking exposures from this regression are then used in the second stage regression with costs as the outcome. The F-statistic from the first stage regression tests the joint significance of all instruments and is an indicator of instrument strength. Weak instruments will lead to estimates biased in the direction of the conventional multivariable estimate.

For the 2SLS models, we developed polygenic risk scores (PRSs), also known as genetic risk scores or allele scores. Each PRS was calculated as the weighted sum of the effect alleles for all SNPs. Each SNP was weighted by the regression coefficient from the respective GWAS in which the SNP was identified. These estimates were adjusted with age, sex and the first 40 genetic principal components.

The interpretation of the 2SLS effect estimates is that of a unit change in genetic liability to the exposure variable. For the initiation exposure, this reflects a change in genetic liability in case status; that is, the causal effect estimate represents the costs per person, per year of becoming a smoker. For the composite index of lifetime smoking, the 2SLS reflects a unit change in the composite smoking index, equivalent to (for example) a current smoker who has smoked 22 cigarettes per day for 10 years, or an individual who smoked 30 cigarettes per day for 11 years who quit smoking 5 years ago. See Wootton et al [42] for further details on this variable.

### 3.3 Sensitivity analysis

We conducted several sensitivity analyses. An important potential violation of the requirements for valid instrumental variable analysis in Mendelian Randomization is horizontal pleiotropy, in which a variant is associated with the outcome other than via the exposure of interest. The presence of pleiotropy may be indicated by the presence of heterogeneity in effect estimates across SNPs. This may be formally tested by comparing Cochran’s Q statistic (Formula 7) to the critical values of a chi-squared distribution:

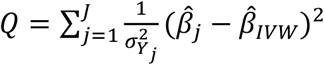

Here, *J* indexes the number of SNPs, 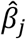 is the effect estimate for SNP *j*, 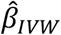 is the overall inverse variance (IVW) weighted effect calculated for *J* SNPs, and the variance of the SNP-outcome association is denoted by 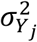. We assessed the sensitivity of our results to potential violations of the exclusion restriction by estimating a variety of over-identified Mendelian randomization models. We implemented a random-effects inverse-variance weighted (IVW) estimator. This estimator calculates the Wald ratio for each SNP separately, and then combines these SNPs using weights determined by the precision of the association between the SNP and healthcare costs. More precisely estimated SNPs receive more weight.

This estimator assumes that no pleiotropy in violation of the exclusion restriction is present, or that any such pleiotropy has a net zero effect on point estimates. The consequence of the latter form of horizontal pleiotropy – in which any effects “balance out” – is that estimates are unbiased but will have somewhat larger variance than in the no-pleiotropy case. We calculated three other estimators that relax this assumption.

The Mendelian Randomization Egger estimator includes and conditions on an intercept, which may be interpreted as the average pleiotropic effect of all included SNPs. Horizontal pleiotropy is indicated if this intercept is statistically different from zero. This estimator is consistent even if all SNPs violate the exclusion restriction, provided further assumptions on the relationship between instrument strength and any direct pleiotropic effect of SNPs are met – for details see [43, 44].

If the Wald ratio estimates for all SNPs are ordered, then the median estimate will be consistent if at least 50% of SNPs are valid instrumental variables. We implemented a penalized and weighted version of a median estimator in which effect estimates are weighted by precision and in which outlying variants (measured by their contribution to the Cochran Q heterogeneity statistic) are penalized by down-weighting their contribution to the overall effect estimate. Finally, the mode estimator is given by the mode of the Wald ratio estimates. Unlike the median estimator, the mode estimator is consistent even if more than 50% of the SNPs are invalid instrumental variables, provided that the largest homogenous cluster of SNPs are valid instrumental variables. We implemented a version of the estimator that down-weighted the contribution of less precisely estimated SNPs.

We also assessed whether genetic variants associated with smoking and smoking heaviness affected costs in “ever smoker” compared to “never smokers” – if these variants influence costs only via smoking heaviness, their effect should only be apparent in ever smokers [45]. We assessed this association by interacting in each of the split GWAS samples the smoking initiation phenotype with variants associated with smoking and located closest to the CHRNA5 locus. Evidence of an interaction in smokers but not non-smokers would constitute some evidence – albeit not definitive evidence – in support of the exclusion restriction for the initiation phenotype.

Smoking SNPs identified in GWASs may also reflect genetic liability to risk tolerance. Risk tolerance may therefore influence smoking behaviour, as well as other risk-taking behaviours that themselves cause healthcare costs. We therefore implemented multivariable Mendelian Randomization, in which the casual effect of both smoking and risk tolerance were jointly estimated in a single instrumental variable model. Multivariable Mendelian Randomization allows for the direct effect (not via the other exposure) of each exposure to be determined. It also allows the causal effect of each exposure to be expressed as conditional on the other. We implemented separate models for smoking initiation and risk tolerance, and for lifetime smoking and risk tolerance. We used the methods of Sanderson and colleagues [46] to implement our models, as implemented in the MVMR R package (https://github.com/WSpiller/MVMR).

We also explored potential consequences of correlated pleiotropy. We applied Steiger filtering [47] to each smoking exposure as a simple test of the assumption that instruments affect the outcome via the influence of these instruments on the exposure. This test calculates the variance explained by the SNPs, and tests if the variance explained by the instrument in the outcome is less than the variance explained in the exposure. If so, this provides some evidence that the SNPs influence the exposure via the outcome, and not because the direction of causation is running from the outcome to the exposure. The weighted median and weighted mode estimators, described above, are robust to some types of correlated pleiotropy, provided that this (and other forms of horizontal pleiotropy) affect less than half of variants (for the median estimator) or that the modal pleiotropic effect is zero (for the modal estimator). We also explored the use of the “MR CAUSE” methodology [48] as a means of accounting for correlated pleiotropy but could not obtain stable estimates.

## 4 Data

We used the UK Biobank study as our primary source of data (13–15). This is a population-based cohort of some 500,000 individuals recruited between 2006 and 2010. All adults aged 40-69 living in defined catchment areas were invited to participate [49, 50], with a final response rate of 5.45%. Participants provided a wide variety of personal and phenotypic data at baseline recruitment, and most consented to genotyping. Individuals in the cohort are generally healthier, wealthier and more educated than the wider UK general population [49, 51].

We studied two measures of smoking: smoking initiation and a composite index of lifetime smoking. The composite index of lifetime smoking was created by Wootton and colleagues [42] to reflect smoking initiation, duration of smoking, heaviness of smoking and smoking cessation (if any). These different measures were aggregated into a composite lifetime smoking index with a half-life constant to reflect the exponential declining impact of smoking on health at a given time. The half-life was obtained from a simulation of the effect of smoking on lung cancer and on all-cause mortality in the UK Biobank.

The smoking initiation phenotype measure captures whether cohort participants ever smoked, without further distinction according to duration or intensity. A binary variable indicating smoking initiation (“ever” smoked versus “never” smoked) was created from the lifetime smoking index, with non-zero values of the lifetime index indicating an individual had initiated smoking.

Mendelian Randomization studies may be biased if the same sample used to select SNPs is also used as the sample in which those SNPs are analyzed as instrumental variables [52]. Sample overlap will tend to bias associations towards the observational exposure-outcome association. We randomly split our available Biobank sample into two non-overlapping samples. On one of these sets, we conducted *de novo* GWASs to identify SNPs, which we then analyzed on the other set of data. We then performed fixed-effect meta-analysis to obtain a single summary measure of effect (and the associated confidence intervals) across both samples. We used a set of standard in-house [53, 54] quality control procedures for conducting the *de novo* UK Biobank GWASs. We used a clumping threshold of R^2^<0·001 to account for potential linkage disequilibrium between SNPs.

Genetic data on lifetime smoking (and the related binary smoking initiation variable) was obtained from two *de novo* GWASs of using the composite lifetime smoking index undertaken on 318,067 individuals in the UK Biobank. We also conducted a *de novo* GWAS for a measure of risk tolerance to use in multivariable Mendelian Randomization (N=274,450). This was constructed using responses to UK Biobank questionnaires. A score was created from response to questions relating to days per week of moderate and vigorous physical activity, hours of TV viewing, breaking of motorway speed limits, illicit drug use, alcohol consumption, self-harm, and sexual activity. The precise details of the construction of this variable are given in the supplementary material.

Genetic and other data from UK Biobank were linked to records of inpatient hospital care. A patient undergoing an inpatient hospital visit will occupy a bed but does not necessarily stay overnight. The process by which episodes of care were coded to reflect costs are described elsewhere [55, 56]. Briefly, episodes of inpatient hospital care were coded to create hospital resource groups (HRGs), which denote episodes of care with similar diagnoses, operations and procedures. These HRGs were then cross-referenced to unit costs of care to create a per-person, per-year overall figure for inpatient hospital costs.

All analyses were performed in *R* version 4.02 and Stata version 16.1. Analysis code is available at www.github.com/pdixon-econ/MR_smoking_costs.

## 5 Results

Up to 300,045 individuals were analyzed, of whom 54% were female (n=161,022). Mean age at recruitment was 57 years (standard deviation = 8.0 years). Mean costs per year were £478 and median costs £87. Some 45% of the sample had zero inpatient NHS costs. There were 87,651 individuals in this sample who had ever smoked. The range of the cumulative smoking index is from 0 to 4, with a score of 0 indicating a never-smoker. The mean value of this index amongst all individuals was 0.33 (standard deviation: 0.67) and amongst smokers 1.14 (standard deviation: 0.78).

Conventional multivariable models, estimated using linear regression and adjusted for age and sex but without any genetic information, are summarized in Table 1.

**Table 1.** Multivariable estimates of association between smoking initiation, lifetime smoking, risk tolerance, and annual inpatient hospital costs.

|  | N | Effect estimate | 95% confidence interval |
| --- | --- | --- | --- |
| **Phenotype** |  |  |  |
| Smoking initiation | 300,045 | £208 | £196 to £219 |
| Lifetime smoking amongst smokers | 87,651 | £142 | £131 to £153 |

Note to Table 1: These models adjust for age, sex and recruitment centre only.

The effect estimate on the smoking initiation phenotype indicates the observational association between becoming a smoker compared to never smoking on per-person, per-year inpatient hospital costs. Likewise, a unit change in the lifetime smoking index in the intensity, duration and cessation (if any) is associated with £142 increase in annual per-person inpatient costs. Given that median per-person costs in this sample are £87 per year, both smoking exposures are associated with a substantial increase in annual costs. These conventional multivariable models will be confounded by any variables other than age, sex and UK Biobank recruitment centre that jointly influence smoking and healthcare costs.

Table 2 details information relating to the polygenic instrumental variable models.

**Table 2.** Cases, number of SNPs and strength of instruments.

|  | N | N of SNPs | % of variance explained by polygenic risk score | F-statistic from first stage of 2SLS polygenic risk score 2SLS model |
| --- | --- | --- | --- | --- |
| **Phenotype** |  |  |  |  |
| Smoking initiation sample 1 | 149,995 | 10 | 0.19% | 223 |
| Smoking initiation sample 2 | 150,050 | 11 | 0.25% | 293 |
| Lifetime smoking sample 1 | 149,995 | 17 | 0.40% | 584 |
| Lifetime smoking sample 2 | 150,050 | 15 | 0.44% | 620 |

Note to table : Percentage of variance obtained from pseudo $R^2$ from a logistic regression of initiation status on the PRS for initiation, and from $R^2$ obtained from linear regression of lifetime smoking on the PRS for this exposure.

The number of SNPs refers to genome-wide significant hits in each split sample of the UK Biobank cohort. The proportion of variance explained by the polygenic risks scores is modest, being less than 1% in all cases. However, the first stage F-statistic from the 2SLS indicates that these are strong instruments.

### 5.1 Instrumental variable results

Table 3 summarises the causal effect estimates and associated 95% confidence intervals representing the effect of genetic liability to each smoking phenotype, estimated using polygenic risk score 2SLS models.

**Table 3.** 2SLS allele score estimates.

|  | Beta | 95% confidence interval |
| --- | --- | --- |
| **Phenotype** |  |  |
| Smoking initiation | £477 | £187 to £766 |
| Lifetime smoking | £204 | £105 to £303 |

Note to Table 3: Estimates and confidence intervals from fixed-effects meta analysis over each split sample.

The betas in Table 3 reflect inpatient hospital costs per person per year of a (genetically influenced) change in liability to smoking initiation and to the lifetime smoking index. Note that these represent the effect of genetic liability to each smoking phenotype, and are not directly comparable to the corresponding conventional multivariable estimates [57, 58]. Note also that the wide confidence intervals are a consequence of the modest proportion of variance that each polygenic risk scores explains in the respective phenotypes. Nevertheless, the effect estimates are consistent with substantial effects of genetic liability to each smoking phenotype on healthcare costs.

### 5.2 Sensitivity analysis

Inspection of Cochran’s Q revealed little evidence of heterogeneity (Table 4).

**Table 4.** Cochran’s Q statistic and heterogeneity by phenotype.

|  | N of SNPs | Q statistic | Q p-value |
| --- | --- | --- | --- |
| **Phenotype** |  |  |  |
| Smoking initiation sample 1 | 10 | 13.2 | 0.16 |
| Smoking initiation sample 2 | 11 | 8.7 | 0.47 |
| Lifetime smoking sample 1 | 17 | 25.2 | 0.07 |
| Lifetime smoking sample 2 | 15 | 12.2 | 0.51 |

Note to table: There were insufficient SNPs to calculate the Q statistic for the risk tolerance exposure.

The data do not suggest the presence of pleiotropy, with the possible exception of sample 1 for the lifetime smoking phenotype. We nevertheless report the findings of the various pleiotropy robust estimators in supplementary material. These data are generally concordant in indicating a similarity of causal effects across estimators for both samples and for both smoking phenotypes. They are similar to the 2SLS analysis in indicating a substantial effect of smoking on costs. The results of Steiger filtering tests indicated that the direction of causality was more likely to be from each smoking exposure to outcome rather than vice versa. These results are reported in supplementary material. Interactions between initiation and variants located near the CHRNA5 locus were consistent with the null (see supplementary material).

We used multivariable Mendelian Randomization to examine whether risk tolerance may influence the association between smoking and healthcare costs. Multivariable Mendelian Randomization requires that the instrumental variable assumptions hold jointly for all included exposures. In particular, SNPs must be strongly associated with each exposure, conditional on the other included exposures. Sanderson and colleagues [46] recommend that each exposure included have an F-statistic greater than 10. In the event, the conditional strengths of the SNPs for risk tolerance (but not the two smoking exposures) were too weak to make reliable inferences. Further details of these results, including the F-statistics, are reported in supplementary material.

## 6 Discussion

Conventional multivariable estimates in simple linear models indicated a substantial association of both smoking initiation and the composite smoking index on annual per-patient inpatient costs in the UK Biobank cohort. Mendelian Randomization analyses of both of genetic liability exposures were estimated with more uncertainty, but were consistent with potentially large impacts on hospital costs.

Under the assumption of monotonicity – that the effect of the instrumental variables on each respective exposure is in the same direction for all subjects – these estimates are local average treatment effect estimates that capture lifelong genetic liability to smoke. In the present study, this assumption means for those individuals whose smoking differs by levels of the respective instruments, then the association change in smoking behaviour is in the same direction for all individuals. This assumption may be reasonable although we do not formally test it.

The Mendelian Randomization estimates refer to the impact on costs of a genetic liability to smoking and do not necessarily correspond with the effect estimates that would be obtained from a hypothetical experiment to compel people to smoke (or to compel smoking cessation) and in which healthcare costs are observed over a period of time. Mendelian Randomization estimates do not necessarily comply with the stable unit treatment value assumption (SUTVA) [59]. Our effect estimates relate to the cumulative lifetime impact of a genetic liability to smoke, and cannot necessarily be used to make inferences about the effects of smoking at particular phases of life. This also complicates comparisons between conventional multivariable models and Mendelian Randomization estimates on the liability scale.

Studies using similar smoking exposures (e.g. [60]) have found evidence of pleiotropic effects, particularly in relation to smoking initiation. There was little evidence of heterogeneity associated with pleiotropy in the present study. We further examined estimates from pleiotropy robust estimators, which were similar and suggested that the same causal effect was being identified by each such estimator. However, if risk attitudes (or other variables) confound the association between smoking and healthcare costs, then interventions that address smoking may not necessarily influence healthcare costs [61]. Moreover, confounding the effects of smoking on healthcare cost with those of attitudes toward risk may lead to biased estimates of the external effects of smoking. There was no attenuation of the effects of smoking on healthcare cost when including instruments for risk tolerance, although this finding was not robust to weak instrument bias. We cannot rule out bias arising from associations between risk attitudes and smoking, including via correlated pleiotropy.

We estimated all associations on the UK Biobank cohort, which is a self-selected sample of individuals who are healthier, wealthier and more educated than the wider UK population of adults. This may lead to sample selection bias. Simulation studies [62, 63] suggest that biases other than those attributable to selection, such as violations of the exclusion restriction caused by pleiotropy, may have a greater impact on effect estimates. We were also limited to studying individuals of European (White British) ancestry, and to the analysis of inpatient hospital costs. The relationships we estimate on this ethnicity may not reflect associations between smoking and costs in other ancestral groups.

Mendelian Randomization requires that variants are conditionally independent of local environments. This assumption will be violated if there is clustering of alleles in certain environments, or amongst certain types of individual. In relation to the former, geographical stratification that is not wholly accounted for by genetic principal components is a feature of UK Biobank [64] and may impart bias to our results. In relation to the latter, alleles that indicate genetic liability to smoking will tend to cluster amongst individuals if assortative mating means that smokers are more likely to marry other smokers than they are to marry non-smokers [65, 66].

There may also be biases from dynastic effects [67]; for example, children raised by smokers are more likely to smoke independently of the genetic liability that a child has to smoke. Howe et al [68] use a within-family GWAS to control for demographic and indirect genetic effects, such as the impact of non-transmitted parental alleles. These GWAS found smaller effect estimates for smoking than in GWASs of unrelated individuals. This would suggest that the effect estimates in the present study, drawn from GWASs of unrelated individuals, may be upward biased.

## 7 Conclusion

Mendelian Randomization analysis of the causal effect of genetic liability smoking initiation and lifetime smoking indicated a substantial impact of each exposure on annual inpatient hospital costs, although these associations may be inflated by indirect genetic effects and potentially also by risk attitudes. Nevertheless, the costs we attribute to smoking suggest that externalities associated with smoking, and the cost-effectiveness of interventions to prevent smoking initiation and encourage cessation, may be considerable.

## Data Availability

Analysis code is available at www.github.com/pdixon-econ/MR_smoking_costs. Data is available on application to UK Biobank

## Acknowledgments

This research was conducted using the UK Biobank Resource under Application Number 29294 and Application Number 9142.

