## Supplementary material for "The causal effect of cigarette smoking on healthcare costs"

### 1 Further detail on Mendelian Randomization and potential violations of the exclusion restriction

The requirements for valid instrumental variable analysis are precisely the same in Mendelian Randomization analysis as in other contexts where instruments other than genetic variants are used. These requirements can be summarized by the directed acyclic graph of Figure A1.

**Figure A1 Directed acyclic graph of an instrumental variable**

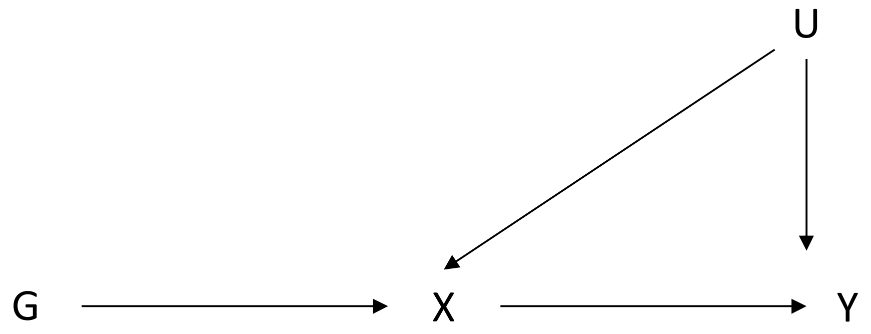


**Notes to Figure 1**: G: genetic variant. X: Exposure variable. Y: outcome variable. U: Confounding variable(s)

Here, G represents a (genetic) instrumental variable, X represents an exposure variable such as smoking initiation or lifetime smoking, and Y represents an outcome. U represents all potential omitted confounding variables, whether known or unknown.

The first instrumental variable requirement is relevance – this requires that instrumental variables be associated with the exposure of interest. This is represented in Figure 1 by the directed graph connecting G to X. Relevance for the genetic variants in Mendelian Randomization analysis is best obtained from replicated GWASs, which measure the association between particular exposures and many – potentially millions – of genetic variants distributed across the genome.

The second variable assumption is conditional independence, sometimes summarized in the phrase “as good as randomly assigned”. In Figure 1, U – representing confounding omitted variables – is connected to both the exposure variable and to the outcome. There is no connection (or directed graph) emanating from the confounding variables U to the instrumental variable G. Since genetic variation is determined at conception, prior to the events, circumstances and behaviors of later life, instrumental variables based on genetic variation are likely to meet this criterion.

However, events that occur prior to conception may conflict with this requirement. For example, sex and year of birth may confound these associations; these influences are typically accounted for by conditioning on sex and age in analysis. More subtle violations of this requirement will occur if the distribution of alleles in a population is correlated with the wider environment. For example, it is known that some allele frequencies differ by ancestry, and this may lead to non-random clusters of genetic variation emerging in particular environments. Conditional independence in these circumstances may be obtained by restricting analyses to individuals of similar ancestry, and by conditioning on genetic principal components.

The third requirement for valid instrumental variable analysis is the exclusion restriction, which requires that the instrument affects the outcome only via its effect on exposure of interest. Paths from the instrument to the outcome that are not mediated by the exposure will violate this requirement. There are two principal mechanisms by which the exclusion restriction may be violated in Mendelian Randomization. These are linkage disequilibrium and pleiotropy, directed acyclic graphs for which are illustrated in Figure A2.

**Figure A2 Linkage disequilibrium and horizontal pleiotropy as violations of the exclusion restriction**

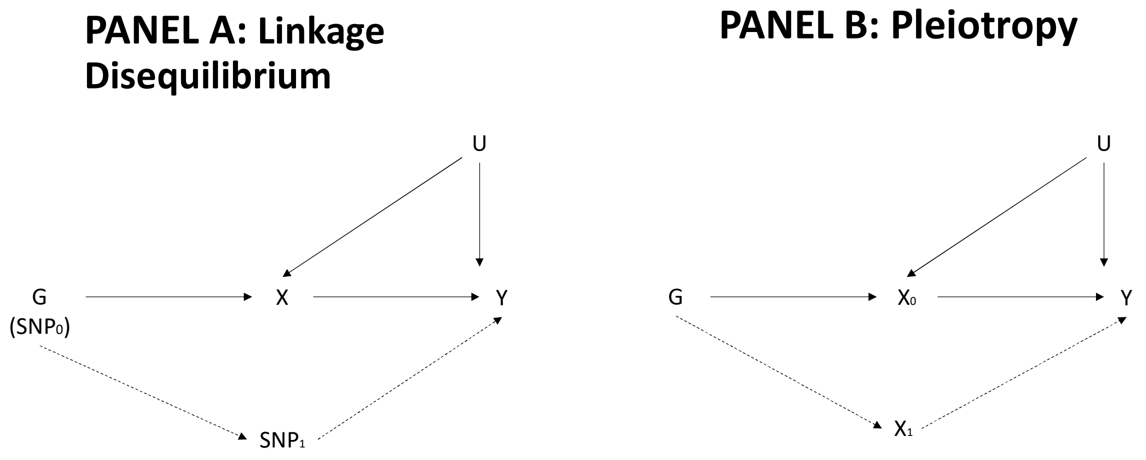


**Notes to Figure 2**: SNP: Single nucleotide polymorphism. G: genetic variant. X: Exposure variable. Y: outcome variable. U: Confounding variable(s)

Linkage disequilibrium refers to fact that genetic variants tend to be inherited together with other variants in close physical proximity on the genome. Including variants in linkage disequilibrium may lead the analyst to incorrectly attribute a causal effect to one of the variants when in truth the causal effect is associated a linked but distinct variant. This may be addressed by restricting analysis to exclude these other variants that are in linkage disequilibrium, typically by filtering ("clumping") based on the strength of the association between variants.

Pleiotropy occurs when one variant affects more than one phenotype. We illustrate horizontal pleiotropy in Panel B of Figure 3, indicating an exposure-independent path from the instrument to the outcome that will violate the exclusion restriction. If the pleiotropy affects a variant that is intermediate between the instrument and the outcome (a situation referred to as *vertical* pleiotropy), or if the other variant does not affect the outcome, then the exclusion restriction will not be violated.

Conditioning on known or potential vertical pleiotropic effects is unlikely to remove all potential biases since not all sources of pleiotropy have been identified. A more robust approach is to test for the presence of pleiotropy and then conduct sensitivity analyses to assess if conclusions alter under different assumptions concerning how the exclusion restriction might be violated.

Figure A3 is a directed acyclic graph representing correlated pleiotropy. The genetic variant G affects both the exposure and the outcome via a shared and unknown heritable factor C.

**Figure A3 Correlated pleiotropy**

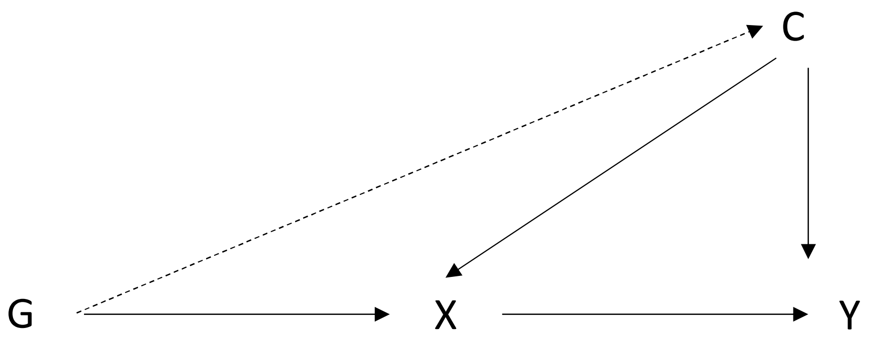


#### 2 Creation of the risk tolerance phenotype

This risk phenotype was created as follows.

- Exercise: Risky behaviour was coded if <5 days per week of moderate or vigorous physical activity

- TV viewing: code >=3 hours per day coded as risky behaviour

- Driving: Ever breaking the motorway speed limit was coded as risky behaviour
- Drug use: Ever using illicit drugs was coded as risky behaviour
- Alcohol consumption: This was coded as risky behaviour if an individual reported drinking daily or almost daily
- Cannabis use: Ever using cannabis was coded as risky behaviour
- Self-harm: Ever engaging in self-harming behaviour was coded as risky behaviour
- Age at first sexual intercourse: Age <16 years at first sexual intercourse was coded as risky behaviour.

Two versions of this score were created. An overall risk score was created by the sum of all responses to these questions indicating a "risky" behaviour, provided that no more than three responses to any of these questions were missing. This is the score reported in the main paper text and refers to the entire sample of individuals (n=274,450 amongst individuals of White British ancestry studied in the main inferential analysis).

The cannabis/drug use/self-harm questions were only asked at online follow up. The second score (approximately n=105,000 were eligible for inclusion in each GWAS) restricted analysis only to those reporting baseline and these online follow-up responses, and with no more than two other categories of risky behaviour missing. For the restricted score, just one SNP was identified in the first split sample, and one for the second split sample, compared to three and two respectively for the larger set. Analysis was re-run on the restricted score – results were similar to those obtained from the unrestricted score, but were even more imprecise than the unrestricted score.

### 3 Pleiotropy-robust sensitivity analysis

This section presents the results of the two-sample summary Mendelian Randomization sensitivity analyses for each smoking exposure in each split sample. Note that the effect estimates are on the scale of the respective genome wide association studies. The composite smoking index was analyzed in its GWAS as a continuous variable on a linear scale. Logistic regression was used to analyze the binary outcome of smoking initiation in each GWAS. The effect estimates for initiation in the tables below therefore correspond to the change in costs per unit change in the log-odds of initiating smoking. A unit change is therefore exp(1), which is 2.72-fold change on the multiplicative scale in the odds of initiating smoking.

**Table A1** **Smoking initiation: Results of summary Mendelian Randomization sensitivity analysis**

| | Estimate | Standard error | P-value |
|---|---|---|---|
| **Smoking initiation sample 1** | | | |
| Inverse variance weighted | £140 | £154 | 0.36 |
| MR Egger | £1,457 | £1,140 | 0.24 |
| Penalized weighted median | £227 | £186 | 0.22 |
| Weighted mode | £214 | £350 | 0.56 |
| **Smoking initiation sample 2** | | | |
| Inverse variance weighted | £402 | £131 | <0.01 |
| MR Egger | £949 | £704 | 0.21 |
| Penalized weighted median | £515 | £179 | <0.01 |
| Weighted mode | £523 | £267 | 0.08 |

**Table A2** **Composite smoking index: Results of summary Mendelian Randomization sensitivity analysis**

| | Estimate | Standard error | P-value |
|---|---|---|---|
| **Composite smoking index sample 1** | | | |
| Inverse variance weighted | £206 | £87 | 0.02 |
| MR Egger | £652 | £391 | 0.12 |
| Penalized weighted median | £191 | £104 | 0.07 |
| Weighted mode | £371 | £218 | 0.11 |
| **Composite smoking index sample 2** | | | |
| Inverse variance weighted | £171 | £73 | 0.02 |
| MR Egger | -£64 | £232 | 0.79 |
| Penalized weighted median | £162 | £103 | 0.12 |
| Weighted mode | £400 | £211 | 0.08 |

The results are consistent with a positive effect of smoking on healthcare costs. Taking into account wide ranges of uncertainty and overlapping confidence intervals around specific point estimates, the results are broadly similar across samples for each exposure, and are broadly similar for the different estimators.

### 4 Results of Steiger filtering

**Table A3** **Steiger filtering results for smoking phenotypes**

| | SNP $R^2$ exposure | SNP $R^2$ outcome | P-value |
|---|---|---|---|
| **Smoking initiation** | | | |
| Sample 1 | 0.003 | <0.001 | <0.001 |

| | | | |
|---|---|---|---|
| Sample 2 | 0.003 | <0.001 | <0.001 |
| **Composite smoking index** | | | |
| Sample 1 | 0.004 | <0.001 | <0.001 |
| Sample 2 | 0.004 | <0.001 | <0.001 |

### 5 Results of smoking initiation interaction test

In each split GWAS sample, we identified SNPs on chromosome 15 and close to the CHRNA5 locus for which their strongest associations in previous studies reported in publicly available databases (including dbSNP and MR Base) were with smoking phenotypes. In sample 1, we used the rs7173514 SNP and in sample 2 rs28681284. For each sample, we estimated linear regressions of the cost outcome on each respective SNP, the smoking phenotype, the interaction of the SNP with the smoking initiation phenotype, and controls for age, sex, UK Biobank assessment centre, and the first forty principal components. In each case, the interaction was consistent with the null. For sample 1, the effect of the interaction was -£11.92 (95% confidence interval -£34.61 to £10.77), and for sample 2 £22.41 (95% confidence interval: -£1.32 to £46.14).

### 6 Multivariable Mendelian Randomization analysis

#### 6.1 A directed acyclic graph for multivariable Mendelian Randomization

A representation of the multivariable Mendelian Randomization model (based on [1]) for two exposures ($X_1$ to $X_2$) is shown in Figure A4.

**Figure A4 Multivariable Mendelian Randomization**

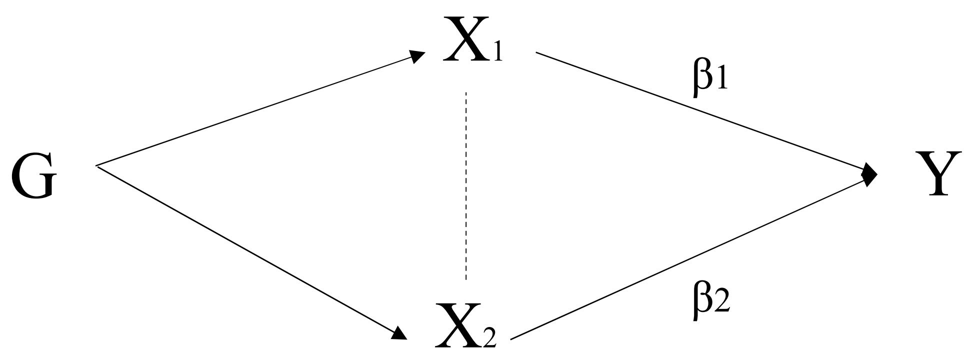

Here, G is a set of SNPs that influences both exposures. In a conventional or univariable Mendelian Randomization analysis, the causal effect estimate reflects the "total" effect of the exposure on an outcome. In a multivariable Mendelian Randomization, the effect estimate for each exposure represents a "direct" effect (respectively $\beta_1$ and $\beta_2$) on the outcome. The sum of the direct effects may not be the same as the total effect, since this depends on the nature of the association between the exposures [1]. This is indicated by the dashed line from $X_1$ to $X_2$.

The model estimated is of the form:

$$Y = \alpha_0 + \beta_1 X_1 + \beta_2 X_2 + \upsilon_y$$

Here, Y is the outcome, $\alpha_0$ is an intercept term, X1 and X2 are exposures that in our analysis represent smoking and risk tolerance. This equation is estimated using two-stage least squares regression, where the first stage regression predicts both exposures from the full set of SNPs that relate to each exposure.

#### 6.2 Results of multivariable Mendelian Randomization

Three SNPs were genome-wide significant in the first split sample (N=129,864) for this phenotype, and just two SNPs the second split sample (N=129,660). The polygenic risk scores created from these SNPs explained 0.10% and 0.05% in the first and second samples respectively. These SNPs were associated with first-stage 2SLS F-statistics of 39 and 28.

The F-statistic for risk tolerance in a multivariable Mendelian Randomization with smoking initiation was 6.1 in the first sample, and 3.7 in the second, compared to 15.0 and 19.3 for the smoking initiation exposure in each respective sample. The F-statistic for risk tolerance in a multivariable Mendelian Randomization with the continuous lifetime smoking index was 7.1 in the first sample, and 4.0 in the second, compared to 20.2 and 27.7 for the smoking index in each respective sample.

Table A4 summarizes the results of the multivariable Mendelian Randomization analysis for each smoking exposure and for each sample.

**Table A4 Results of multivariable Mendelian Randomization**

| | Estimate | Standard error | P-value |
|---|---|---|---|
| **Initiation** | | | |
| Smoking initiation sample 1 | £272 | £126 | 0.04 |
| Risk tolerance sample 1 | -£118 | £93 | 0.21 |

| | | | |
|---|---|---|---|
| Smoking initiation sample 2 | £381 | £95 | <0.01 |
| Risk tolerance sample 2 | -£68 | £112 | 0.54 |
| **Composite smoking index** | | | |
| Composite smoking index sample 1 | £198 | £76 | 0.02 |
| Risk tolerance sample 1 | -£118 | £86 | 0.18 |
| Composite smoking index sample 2 | £224 | £61 | <0.01 |
| Risk tolerance sample 2 | -£29 | £112 | 0.80 |

The effect estimates for the smoking exposures may be compared to the inverse variance weighted estimates in Tables A1 and A2. These estimates are broadly similar in that analysis and the multivariable analysis reported in Table A4, indicating that conditioning on instruments for risk tolerance does not appear to materially alter the impact of smoking on healthcare cost. We emphasize that the results are not robust to possible weak instrument bias (as noted in the main text) and are reported here for completeness.

### 7 Supplementary material references
